# Data Resource Profile: the Physical Activity Cohort Repository (PACe)

**DOI:** 10.1101/2025.07.22.25332023

**Authors:** Leonessa Boing, Andrea Ramirez Varela, Saud Abdulaziz M Alomairah, Derrick A. Bennett, Lucimére Bohn, Clare Hume, Elli Kontostoli, Sigríður Lára, Paul Mackie, Jacqueline Louise Mair, Clarice Martins, Jorge Mota, Andreia Pizarro, Jenny Rossen, Jakob Tarp, Kirstie Tew, Philip von Rosen, Terry Boyle, Brigid M. Lynch

## Abstract

**Key Features:**

* The Physical Activity Cohort Repository (PACe) project created a searchable online database of cohort studies that have prospectively collected data on physical activity and/or sedentary behaviour (self-report and/or device-based measurement) at three or more timepoints. Only cohorts with ≥ 1,000 participants at baseline were included.
* The PACe is a freely available resource created to encourage researchers to return to existing cohorts to apply contemporary causal inference methods that appropriately deal with time varying exposures and confounders, to improve the quality of evidence pertaining to physical activity, sedentary behaviour and health.
* Two hundred and nineteen unique cohorts were included in the initial iteration of the PACe, representing all World Health Organization regions. The earliest included cohort started in 1922 (the Terman Life-Cycle Study) and extends to ongoing cohorts.
* The development of the PACe was an international effort; this resource was designed to facilitate global collaboration and building research capacity in low- and middle-income countries.
* We will update the repository every two years, to ensure that new cohorts and cohorts adding additional waves of follow-up are captured.

## Data resource basics

The Physical Activity Cohort Repository (PACe) project was first proposed by the Epidemiology Council of International Society of Physical Activity (ISPAH) in March 2019. The project was approved by the ISPAH Board, volunteer members from the Epidemiology Council were recruited, and the project commenced in June 2020.

The project aimed to create a central database of cohort studies that have prospectively collected data on physical activity and/or sedentary behaviour (self-report and/or device-based measurement) at three or more timepoints, and include at least 1,000 participants at baseline.

Critical assessment of the epidemiological methods used to measure physical activity and sedentary behaviour variables in longitudinal studies is necessary continue building the evidence base pertaining to physical activity, sedentary behaviour and health outcomes.^1^ Yang *et al*. have highlighted the methodological shortcomings of longitudinal studies of physical activity and/or sedentary behaviour and mortality.^2^ Very few studies to date have used appropriate methods to handle time-varying exposures in the presence of time-varying confounders. Selection bias (i.e. collider stratification bias) can occur if a time-varying confounder that shares a common cause with the outcome is adjusted for using conventional regression methods.^2,3^ We therefore purposely selected only cohorts with at least three waves of data collection, to enable researchers to apply contemporary causal inference methods. For example, a researcher interested in emulating a target trial to estimate the effects of sustained physical activity interventions on all-cause mortality would find the PACe a helpful tool to identify cohort studies that have an appropriate data structure to apply g methods.

There is also a need to build physical activity research capacity in low income countries; there exists a scarcity of local evidence in these settings, which also experience high burdens of non-communicable disease.^4^ This repository of cohort studies will help to maximize the investments made into cohort studies by governments and the civil sectors around the world, by encouraging researchers to return to existing studies and facilitating international collaboration.

## PACe location and support

The PACe can be accessed at: https://www.cancervic.org.au/research/epidemiology/pace

The content of the PACe will be updated every two years by the Epidemiology Council of ISPAH. The web interface is supported by the Cancer Epidemiology Division of Cancer Council Victoria, a non-profit cancer charity organisation involved in cancer research, prevention, advocacy and patient support. Cancer Council Victoria maintains the PACe web interface as part of its goal to discover new prevention, detection, support, and treatment opportunities to improve cancer outcomes and save lives. No end date for this support is scheduled.

## Protocol for data collection

The PACe scoping review protocol was published in 2020 on the Epidemiology Council page of the ISPAH website https://ispah.org/wp-content/uploads/2020/05/Protocol-for-ISPAH-website_20200428.pdf.

### Eligibility criteria

The inclusion criteria were: (i) The study must be a prospective cohort study with three or more waves of data collection that include measures of physical activity and/or sedentary behaviour; Physical activity and/or sedentary behaviour measurements can be self-reported, or device based; Physical activity and/or sedentary behaviour may also be retrospectively collected (e.g. adult participants asked about physical activity during childhood), but this retrospective assessment does not count as one of the three required waves of physical activity assessment; and (iv) No restriction on sample size or populations and outcomes.

Where multiple research papers were published from the one cohort study, we used the (i) cohort profile if one exists; (ii) the publication that best describes measures of physical activity and/or sedentary behaviour; and (iii) the publication that describes the maximum number of waves of data collection, as required.

### Data sources and search strategy

The search was implemented in January 2020 in two electronic databases: PubMed and Web of Science. For the main search we used the terms “physical activity” OR “physically active” OR “physical inactivity” OR “physically inactive” OR “fitness” OR “exercise” OR “exercises” OR “walk” OR “walking” OR “sedentary” OR “active transport” OR “active transports” OR “active transit” OR “active travel” OR “commute” OR “active commuting” OR “bicycle” OR “bicycling” OR “bike” OR “biking” OR “active living” OR “active-living” AND “cohort profile” OR “cohort study”. To identify the cohort profiles and complement the first search, a second search was performed on PubMed using the terms “International journal of epidemiology”[Jour] AND “cohort profile”. There was no age, dates of publication or language restrictions.

### Selection of sources of evidence

Fifteen reviewers working in pairs (one group of three) evaluated a portion of the titles and abstracts identified by the search, and made recommendations for full text review. Each team then sequentially reviewed the full texts assigned to their group and extracted data from relevant cohorts. When conflicting decisions was made, other authors were consulted (BL, TB, ARV, LB). Covidence systematic review software was used for screening titles and abstracts, finding duplicates and full-text screening.

### Data extraction

A customised data extraction template was developed using Google forms. Data extracted from the eligible full texts summarised study characteristics in three sections: (i) **cohort characteristics** - link to cohort profile (if applicable) or contact details of Principal Investigator, years of study commencement, country, state or city where the cohort was conducted, country region classification according to the World Health Organization, country income level classification according to the World Bank, sample size at baseline, cohort type (e.g. birth cohort, occupational cohort, other), study population age group (adult population ≥ 18 years < 60 years, children and adolescents < 18 years, specific for older adult population ≥ 60 years, or more than one age group), number of waves of data collection including physical activity and/or sedentary behaviour; (ii) **physical activity and/or sedentary behaviour measurement** - if it was self-reported or objectively assessed, and which questionnaire or instrument was used; and (iii) **other variables measured** - sociodemographic factors, health status during pregnancy (in case of birth cohorts), health status of participants including chronic conditions and comorbidities, anthropometry, sleep, other behaviours (e.g. smoking, dietary intake, alcohol), clinical examination (e.g. blood pressure, heart rate), biological samples collected (e.g. blood, saliva), neighbourhood characteristics, geo-coding, mortality via vital status linkage, cancer incidence via cancer registry linkage, and other disease endpoints via administrative data linkage (e.g. hospital admissions).

### Risk of bias

Risk of bias was not conducted for this scoping review, primarily because the purpose was to identify cohort studies with measures of physical activity and/or sedentary behaviour to create the PACe as a research resource for ISPAH members. Each unique cohort will have its own biases to be considered by researchers utilising data for secondary analyses.

## Results

The search results are presented in Figure 1. The initial data search identified 10,750 papers, of which 7,993 studies were from PubMed and 3,011 from Web of Science. A second search in PubMed focused only on cohort profiles published in the International Journal of Epidemiology and identified 437 cohort profiles. After duplicates were removed 8,992 studies were included in title and abstract screening. Another 6,746 studies did not meet our inclusion criteria were excluded, leaving 2,286 papers for full text screening. A total of 218 unique cohort studies met the PACe inclusion criteria.

**FIGURE 1:**
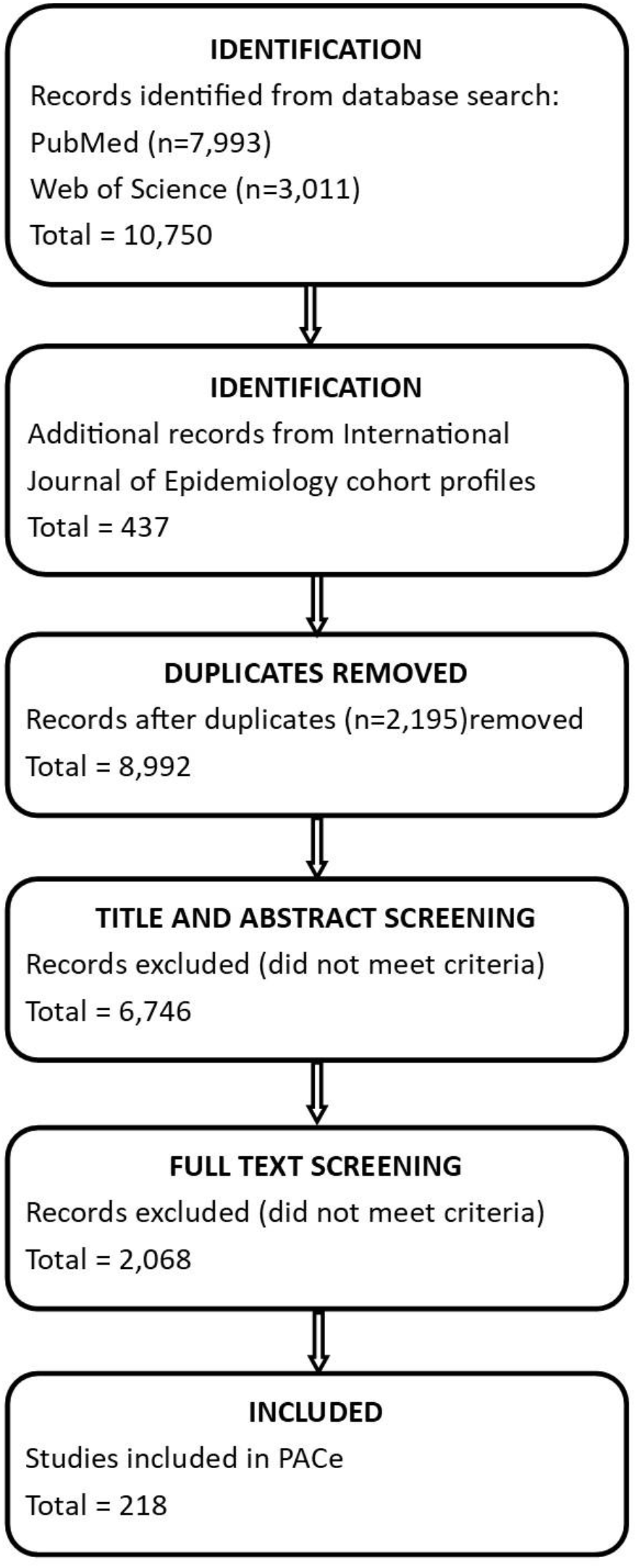
PRISMA flow diagram incorporating literature search, screening and study selection.

Key characteristics were selected to feature as selection criteria on the web interface (see Figure 2). Firstly, if the cohort name is known, the user can find and select the cohort then download the total summary information as a csv file. If the user wants to search across the PACe platform to identify potential studies for their research project, the following search filters can be applied: WHO region, country income level, cohort type, type of physical activity measurement, type of sedentary behaviour measurement, disease endpoints, and whether biological samples were collected. Users can employ some or all of the search filters, depending on how broad or narrow they wish to make their search. The PACe will list all cohorts meeting the defined criteria, for review online or for downloading as a csv file that will include all extracted data for each cohort.

**FIGURE 2:**
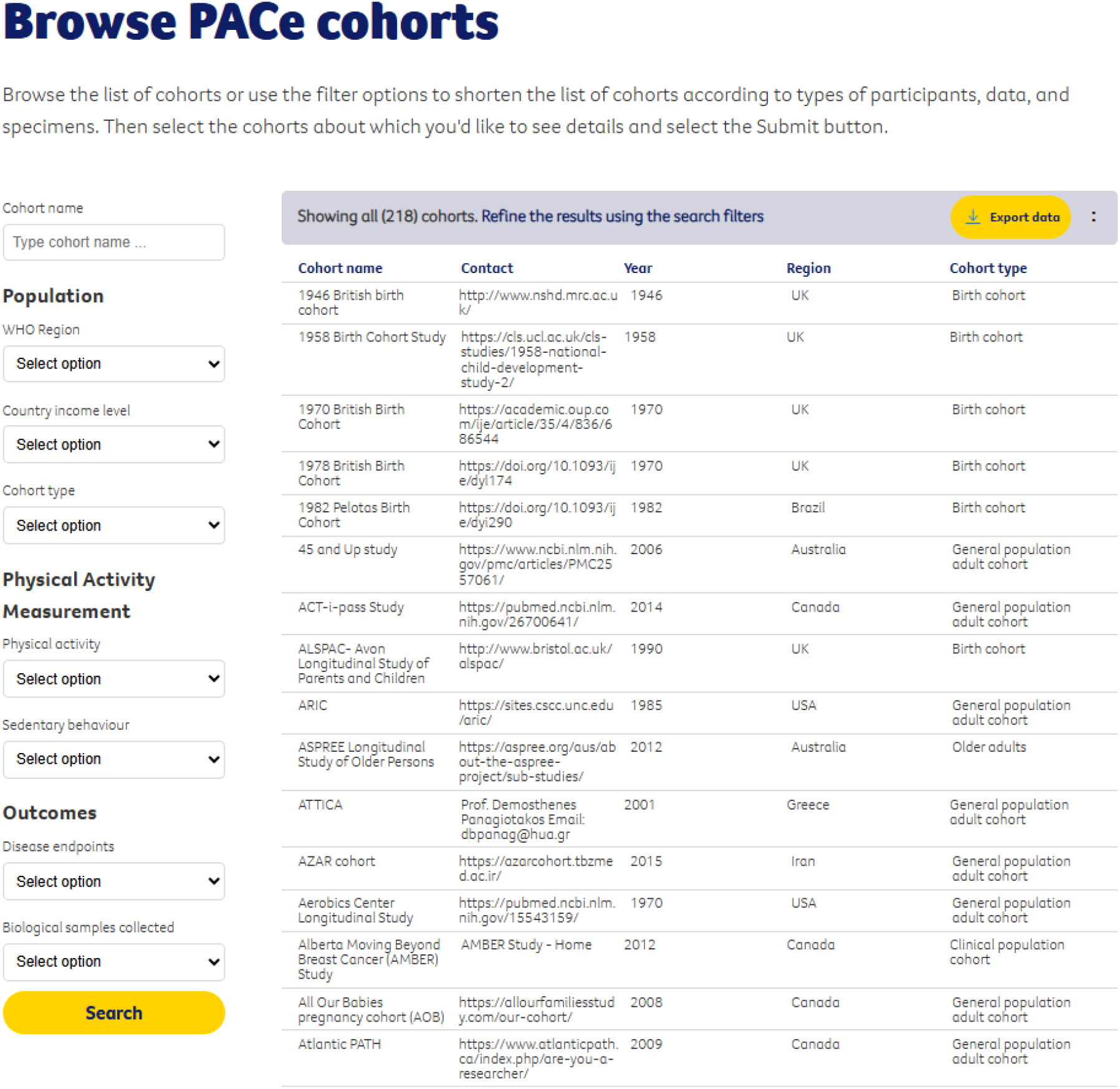
PACe web interface with search filters.

## Data resource use

### Example 1: Scoping review of objective assessment of physical activity worldwide

The Global Observatory of Physical Activity (GoPA!) utilised the PACe to help identify observational studies of adults that used device-based measurement of physical activity (accelerometers, motion sensors, heart rate monitoring).^5^ Forty-two eligible cohorts were identified using the PACe. The scoping review found that objective physical activity data were available for 36 countries. Europe and North American were over-represented, whilst there were only three studies from Africa and no studies from the Eastern Mediterranean or South-East Asia. No meaningful differences were observed between high income, upper middle income and lower middle income countries.^5^

### Example 2: Identifying cohorts to estimate mortality effects of hypothetical interventions on physical activity

The vast majority of studies on physical activity and mortality have used exposures and confounders measured at a single time point, and therefore do not assess the effects of exposure changes over time. Conventional regression modelling can be problematic when there is time-varying confounding that is influenced by prior exposures (e.g. physical activity affects adiposity, which in turn affects physical activity at the next time point).^2,6^ One method that can deal with both time-varying exposures and confounders is the parametric g-formula. This method allows for estimation of the causal effects of hypothetical interventions on physical activity in a population, which can be more informative for public health policy than a typical regression model exploring an ‘association’.

The PACe is an ideal place to identify cohorts that have three or more measures of physical activity and its confounders, along with follow-up vital status data, to facilitate addressing the research question posed for Example 2. With the PACe web interface set to cohort type of “General population adult cohort”, physical activity measurement of “Self-report only” and disease endpoint of “Mortality” we identified 58 cohort studies worldwide that have at least three exposure timepoints.

## Strengths and weaknesses

Despite undertaking a comprehensive systematic review to identify cohort studies eligible for inclusion in the PACe, it is possible that there are studies that have been missed. The corresponding author of this resource profile can be contacted by cohort investigators or other researchers who identify a missing cohort. We are in the process of updating the systematic review to include cohorts that have been established and/or achieved three time points of physical activity or sedentary behaviour exposure assessment since the initial search. Cohorts will continue to be added to the repository for the foreseeable future.

Strengths of this data resource include being freely available via a web browser, no requirement for any special software or coding skills, and that it is maintained by a long-standing and highly respected not-for-profit organisation (Cancer Council Victoria).

## Data resource access

The freely available Physical Activity Cohort Repository (PACe) can be found at: https://www.cancervic.org.au/research/epidemiology/pace. To report errors or missing cohort studies please contact.

The PACe provides the website for each included cohort study (where information regarding data access requests can be found) or the email address for the study Principal Investigator if the cohort does not have its own website. Users of the PACe must make their own application for data access directly with the cohort studies.

## Data Availability

All data produced are available online at https://www.cancervic.org.au/research/epidemiology/pace

## Author contributions

LB, ARV, TV and BML led the conceptualisation of the Physical Activity Cohort Repository (PACe). LB and BML had operational oversight of the project. All authors contributed to screening and data extraction via the systematic review. LB and BML drafted the manuscript text. All authors contributed to testing of the beta version of the web interface. All authors provided an in-depth review of the manuscript and approved the final version.

## Acknowledgements

We thank Bhavika Patel from Cancer Council Victoria for developing the PACe website and user interface.

